# Development and validation of a tool to appraise guidelines on SARS-CoV-2 infection prevention strategies in healthcare workers

**DOI:** 10.1101/2020.06.14.20130682

**Authors:** Ashwin Subramaniam, Mallikarjuna Reddy, Alexander Zubarev, Umesh Kadam, Zhengjie Lim, Chris Anstey, Shailesh Bihari, Jumana HAji, Subhathra Karunanithi, Kollengode Ramanathan, Jinghang Luo, Neil Mara, Saikat Mitra, Arvind Rajamani, Francesca Rubulotta, Erik Svensk, Kiran Shekar

**Author notes:** **Corresponding Author:** Dr Ashwin Subramaniam, MBBS, MMed, FRACP, FCICM, Intensive Care Specialist, Frankston Hospital, 2 Hastings Road, Frankston, VIC, Australia 3199.

## Abstract

**Background:** Clinical guidelines on infection prevention strategies in healthcare workers (HCWs) play an important role in protecting them during the SARS-CoV-2 pandemic. Poorly constructed guidelines that are not comprehensive and are ambiguous may compromise HCWs’ safety. We aimed to develop and validate a tool to appraise guidelines on infection prevention strategies in HCWs.

**Methods:** A 3-stage, web-based, Delphi consensus-building process among a panel of diverse HCWs and healthcare managers was utilised. We validated the tool by appraising 40 international, specialty-specific and procedure-specific guidelines along with national guidelines from countries with a wide range of gross national income.

**Results:** Overall consensus (≥75%) was reached at the end of three rounds for all six domains included in the tool. The chosen domains allowed appraisal of guidelines in relation to general characteristics (domain-1), recommendations on engineering (domain-2) and administrative aspects (domain 4-6) of infection prevention, as well as personal protection equipment (PPE) use (domain-3). The appraisal tool performed well across all domains and inter-rater agreement was excellent. All included guidelines performed relatively better in domains 1-3 compared with domains 4-6 and this was more evident in guidelines originating from lower income countries.

**Conclusion:** The guideline appraisal tool was robust and easy to use. Recommendations on engineering aspects of infection prevention, administrative measures that promote optimal PPE use and HCW wellbeing were generally lacking in assessed guidelines. This tool may enable health systems to adopt high quality HCW infection prevention guidelines during SARS-CoV-2 pandemic and may also provide a framework for future guideline development.

**Funding:** No funding received.

**Key Summary:** We developed and validated a guideline-appraisal tool by appraising 40 different guidelines from countries with varying GNI. This tool may help healthcare systems to adopt high-quality HCW infection-prevention guidelines during COVID-19 pandemic and may also provide a guideline development framework.

## Introduction

The highly transmissible Severe Acute Respiratory Syndrome Coronavirus 2 (SARS-CoV-2), as of 28^th^ of May, has infected more than 5.8 million worldwide [1] and has imposed an unprecedented burden on healthcare systems globally [2]. A significant proportion of the infections recorded during the global pandemic have occurred in healthcare workers (HCWs) [3-6], with many dying as a result [7-10]. Accurate data on prevalence and outcomes of SARS-CoV-2 infection in HCWs around the world is lacking. Prevention of the exposure to the virus is the cornerstone of safe practice for HCWs involved in care of SARS-CoV-2 infected hospitalized patients with coronavirus disease 2019 (COVID-19). This can be achieved by engineering solutions that are designed to minimize the risk of exposure, building administrative processes that alter work practices and through optimal use of personal protective equipment (PPE). Risk of contracting COVID-19 increases in the absence of effective PPE, suboptimal training in the correct use of PPE, and reusing or fashioning own PPE out of inappropriate materials [4][11, 12][13]. This is particularly relevant when health services are experiencing a state of surge.

During the pandemic from a novel virus, health systems and HCWs have to rapidly adopt various infection control strategies such as engineering, administrative and PPE soltuions to create a safe work enivronment and protetct HCWs. Well-designed infection control guidelines based on available evidence, previous experience and expert opinion can play a signfiacnt role in expedited re-organization of the healthcare services creating a safe work environment. The first guideline, based on rapid advice WHO guideline development methods, released soon after the initial COVID-19 outbreak in Wuhan, strongly recommended appropriate protection for all HCWs caring for patients with COVID-19 illness [14]. Various guidelines have since been published in quick succession due to the apparent need for direction in these uncertain times [15].

HCW infection rates vary between countries and the reasons behind this are probably multifactorial and are not entirely clear. Robust specific guidelines on SARS-CoV-2 infection control measures in healthcare facilities can help to ensure high standards of patients and HCWs safety in line with the best available evidence on clinical care and cost effectiveness. However, developing universal international infection control standards may be challenging and not always possible given the disparities in socioeconomic conditions and health care infrastructure around the world. This may be reflected in the recommendations made in the published infection control guidelines. Therefore, we aimed to develop a new consensus-based tool for appraisal guidelines on COVID-19 infection control in healthcare facilities. In addition, we aimed to validate the tool by testing it on various international and national, generic and specialty-specific guidelines available at the time that address the issue.

## Methods

This research did not include collection or distribution of any sensitive medical information and hence ethics approval was not required.

### Development of a PPE guideline appraisal tool

#### The Delphi Panel

The purpose of the Delphi panel was to reach consensus on the guideline appraisal tool [16]. The panel included 84 participants (that also included 17 authors, Supplementary table 2) and comprised of medical managers, intensive care specialists, anaesthetists, infectious disease specialists, intensive care nurses and educators, infection control nurses, emergency, respiratory and general physicians, surgeons (including ear nose and throat specialists and dentists), general practitioners, hospital executives, junior doctors, patient services assistant and data managers. The details of the Delphi process are summarised in Figure 1. Input from the panel was obtained using a 3-step process. Each step comprised of a web-based survey, the results of which were discussed in web-based meetings, followed by real-time polling of participants of the web meeting.

**Table 2:** Agreement scores between Domains with reliability coefficients. Results indicate very good agreement between the assessors. The Delphi system used was reliable and robust with excellent inter-rater agreement.

| Domain | Assessor 1 | Assessor 2 | p-value | Spearman's rho (ρ) | Cronbach's alpha (ρ<sub>T</sub>) |
| --- | --- | --- | --- | --- | --- |
| 1 | 6 (5, 7) | 6 (5, 7) | 0.491 | 0.60 | 0.78 |
| 2 | 5 (4, 7) | 5 (2, 7) | 0.210 | 0.71 | 0.84 |
| 3 | 5 (1, 7) | 4 (1, 7) | 0.198 | 0.73 | 0.86 |
| 4 | 4 (1, 7) | 3 (1, 6) | 0.047 | 0.73 | 0.85 |
| 5 | 1 (1, 5) | 2 (1, 4) | 0.904 | 0.72 | 0.83 |
| 6 | 1 (1, 1) | 1 (1, 1) | 0.965 | 0.65 | 0.86 |
| All |  |  |  | 0.79 | 0.88 |
| Spearman's rho assesses monotonicity to assess whether the two variables rise and fall together. Cronbach's alpha is a commonly used score that measures agreement between the two variables. |  |  |  |  |  |

**Figure 1:**
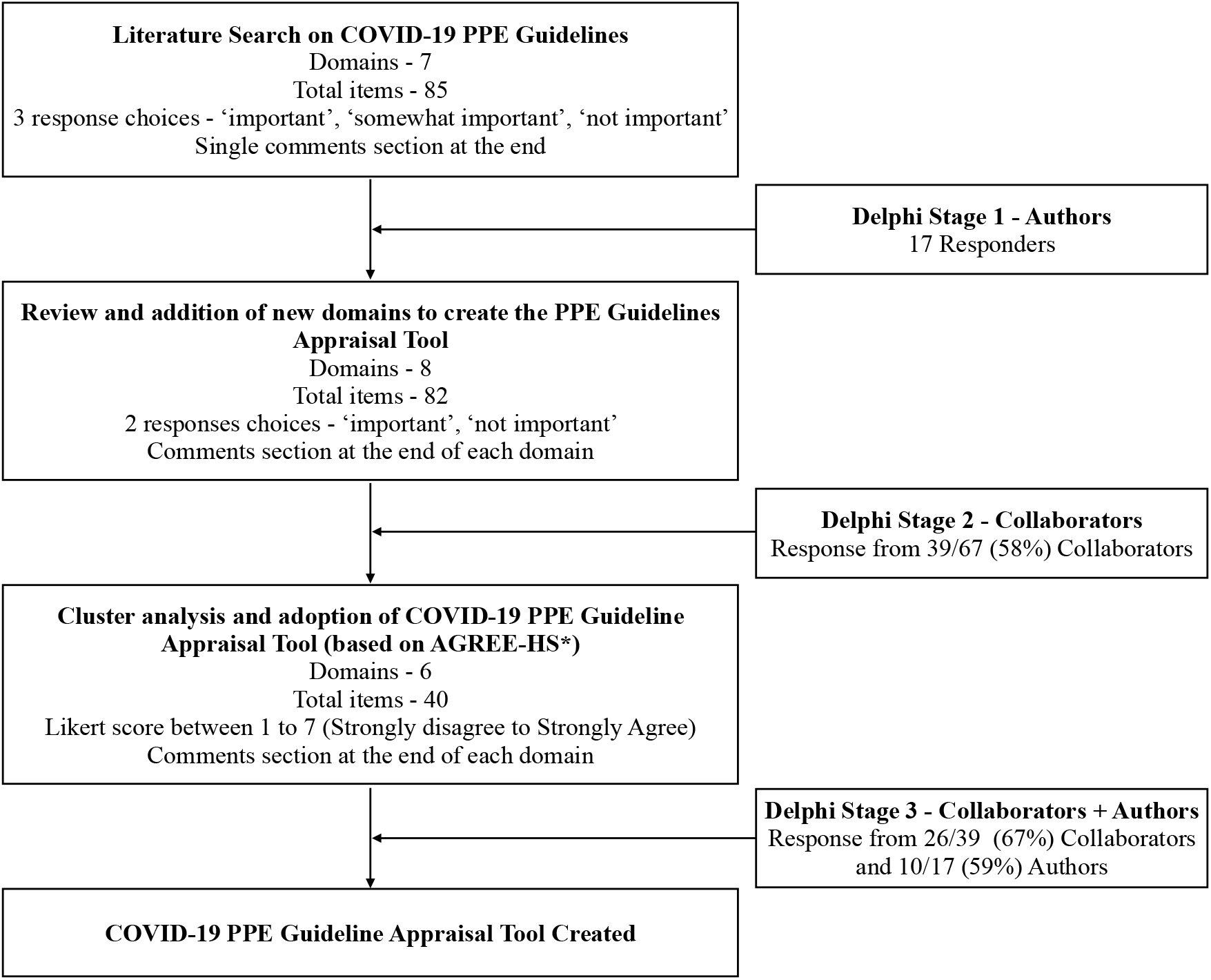
Flowchart of the Delphi survey showing the number of participants, and number of tools and domains from stages 1 to 3. Collaborators were invited to participate in Delphi 2 and 3 surveys. 39/67 (58%) participants invited for Delphi 2 completed the survey. Delphi 3 survey was sent to those 39 collaborators who completed Delphi 2 along with authors of the study. 26/39 (67%) collaborators and 10/17 (59%) authors completed Delphi 3 survey.

#### Delphi Tool construction and reduction of the Appraisal tool

Using a modified delphi process, we developed an initial survey after literature review and web-based discussions between authors. Authors participated in delphi 1 survey and collaborators were invited to participate in delphi 2 and 3 surveys. The 7-domain, 85-item delphi round 1 questionnaire (Supplementary Table 1) expected the respondents to mark one response out of ‘very important’, ‘somewhat important’ and ‘not important’. Based on the respondents’ feedback, a refined round 2 survey with 8 domains and 82 items was constructed (Supplementary Table 2). In delphi round 2, the panel members were requested to mark the items on a two-point Likert scale, either as “Important” or “Not Important”. After the second round, the survey was refined to six domains comprising 40 items. In round 3, for each of the 6 domains, participants were asked to rate the importance of assessing the domains in infection control guidelines using a 7-point Likert scale: 1 = strongly disagree, 2 = disagree, 3 = mildly disagree, 4 = neutral, 5 = mildly agree, 6 = agree and 7= strongly agree. Participants were explicitly instructed to evaluate the general concept of each domain. We did not offer any particular instrument used to measure these domains (Figure 1). We analysed the results of the third round for agreement and degree of consensus. Only data for participants who completed both rounds were included in the results. Consensus was defined as a minimum average score of 5.25 (75%), meaning that the Delphi process would continue until at least 75% of the panel agreed a component should be included in the final tool [17].

**Table 1:** Breakdown of individual Domain scores for Delphi 3

| **Domains** | **Mean (SD)**<br>**Score** |
| --- | --- |
| **Domain 1** |  |
| 1. Guideline specific to COVID-19 | 6.42 (1.30) |
| 2. Targeted at broad range of healthcare workers | 5.83 (1.38) |
| 3. Guideline based on robust evidence/best evidence where available | 6.50 (1.16) |
| 4. Layout of document (Clarity, use of tables, animation, pictures) | 6.17 (1.21) |
| 5. Guidelines must be easy to follow | 6.58 (0.87) |
| **Domain 2** |  |
| 1. Guideline should recommend dedicated areas for safe patient care within the hospital | 6.11 (1.04) |
| 2. Guideline recommends PPE in context of these areas of care | 6.55 (0.91) |
| 3. Guidelines should discuss isolation rooms (e.g. being negative pressure, negative flow or positive pressure) | 6.44 (0.97) |
| 4. Provides recommendations in relation on appropriate areas for performing AGPs | 6.53 (1.00) |
| 5. Dedicated and separated areas for donning and doffing | 6.28 (1.03) |
| **Domain 3** |  |
| 1. Equipment specific for droplet precautions | 6.36 (1.13) |
| 2. Equipment for performing aerosol generating procedures | 6.69 (0.75) |
| 3. Equipment specific for emergency situations (MET, code blue, cardiac arrest) | 6.53 (0.91) |
| 4. Equipment specific for intra-hospital transfer | 6.28 (1.21) |
| 5. Equipment specific for inter-hospital transfer | 6.00 (1.41) |
| **Domain 4** |  |
| 1. Hand hygiene | 6.28 (1.26) |
| 2. Recommends regular training for donning / doffing | 6.44 (1.21) |
| 3. N95/P2 Fit test | 6.17 (1.18) |
| 4. Buddy system always present | 6.17 (1.40) |
| 5. N95/P2 Fit check | 6.58 (0.84) |
| 6. Recommends regular training for PPE use for AGPs | 6.42 (1.08) |
| 7. Clinical disposal for doffed PPE (biohazard waste) | 6.39 (1.05) |
| 8. Recommends training for cleaners | 6.34 (1.22) |
| **Domain 5** |  |
| 1. Minimum mandatory use of PPE | 6.47 (1.23) |
| 2. Duration of use of a single PPE | 6.47 (0.84) |
| 3. Prioritisation of PPE for non-ventilated patients (on HFNO / NIV) | 6.22 (1.02) |
| 4. Prioritisation of PPE for Airborne precaution | 6.44 (0.91) |
| 5. Prioritisation of PPE for AGPs | 6.95 (0.75) |
| 6. Prioritisation of PPE for mechanically ventilated patients | 6.00 (1.22) |
| 7. Recommendations provided for PPE re-use | 6.14 (1.36) |
| 8. Recommendations on how to sterilise PPE prior to re-use N95 and/or face shield | 6.03 (1.48) |
| **Domain 6** |  |
| 1. Incident reporting systems for breaches in PPE /Infection control | 5.44 (1.52) |
| 2. Post-exposure management | 6.28 (1.19) |
| 3. Psychological health support and well-being for all essential staff and health care workers with the care and management of COVID-19 patients | 5.81 (1.62) |
| 4. Staffing and fatigue policies | 5.89 (1.35) |
| 5. Staff amenities meals, rest areas | 5.86 (1.31) |
| 6. Recommendations for Healthcare worker at high risk | 6.28 (1.03) |
| 7. Accommodation for staff on active COVID duty to avoid going home to family | 5.58 (1.31) |
| 8. Length of COVID duty in each shift (4/6/12 hours) | 5.75 (1.50) |
| 9. Guidance on post-COVID duty/shift precautions | 5.78 (1.49) |
| COVID-19 Coronavirus disease 2019, MET – medical emergency team, AGP -aerosol generating procedure, PPE – personal protective equipment, HFNO – high flow nasal oxygenation, NIV – non-invasive ventilation |  |

### Validation of the PPE Guideline Appraisal tool

We conducted a literature search using Pubmed, Embase, Web of Science, The Cochrane Central Register of Controlled Trials (CENTRAL), and CINAHL using the keywords “COVID-19” and “Guidelines” or “Recommendations” in the title which identified 33 and 59 articles respectively (Supplementary Figure 1). National COVID-19 specific guidelines that were not published in medical journals were found on the internet using Google and Bing search engines or obtained from the authors’ professional contacts in these countries by personal correspondence. No language restrictions were placed during the search. All guidelines published in foreign languages were translated to English by the survey participants. After a thorough selection process, we chose to appraise 40 guidelines published between December 1^st^, 2019 to April 30^th^, 2020 (Table 1): This included four international guidelines (G24 and G39, both WHO Guidelines; G27, European Centre for disease prevention and control; and G36, European International Liaison Committee for Resuscitation) and 23 national guidelines from each of high-income (n=9, G1-G9), upper middle (n=4, G10-13), lower middle (n=6, G14-G18, G22) and low-income (n=4, G19-12, G23) countries. The selection of national guidelines was primarily based on the country classification by Gross National Income (GNI) per capita levels.[18] In addition, specialty-specific (n=8, G25-G32; emergency, critical care and anaesthesia) and procedure-specific (n=8, G33-G40; intubation, cardiopulmonary resuscitation and handling of the deceased) were also included. Guidelines were allocated for appraisal to nine reviewers who were all experienced clinicians and frontline COVID-19 HCWs. Guidelines allocation was random, regardless of the reviewers’ background or preferences, however a reliable blinding was not feasible for the study. Each of the 40 guidelines were independently appraised by two reviewers, except the WHO guideline, which was appraised by five reviewers. The list and references of the 40 guidelines are provided in supplementary document.

### Statistical analysis

The differences in the Likert scores assigned by the assessors for each of the six domains were recorded for each of the 40 guidelines included in the final analysis. Guidelines (1-23) were tested first using clustering by Domain (1-6) and second clustering by guideline type (25-40; specialty-specific and procedure-specific). Guideline 24 promulgated by the WHO was treated separately in both cases as it was graded by 5 assessors. Descriptive statistics for normally distributed data were presented using the mean and standard deviation (SD), while median and inter-quartile range (IQR) was used to describe non-normally distributed data. Dichotomous and categorical data were described using frequencies and percentages. Normality was assessed using the Shapiro-Wilk test (SWT). The Likert scores each GNI group were normally distributed and illustrated as mean and 95% confidence interval. Binary comparisons were analysed using either a standard t-test for normally distributed data or Wilcoxon’s rank-sum test for non-normal data. If the data were paired, the appropriate paired tests were employed. Group comparisons for categorical data were analysed using Kruskal-Wallis H test. General correlation and monotonicity were analysed using Spearman’s rho (ρ) and inter-rater reliability was analysed using Cronbach’s alpha (ρT). The level of significance was set at α=0.05 throughout. STATA™(version 15.1) was used for all analyses.

## Results

### The Delphi Process

After the first-round survey, the Delphi panel members were invited to participate in the web-based survey rounds two and three that were circulated 3 days apart (Figure 1). The 54/80 (68%) participants who completed round 2 were invited to participate in round 3. Round 3 was completed by 36/59 (61%) of panel members. The demographics of the participants in the Delphi consensus process are summarised in Supplementary Table 3. The round 3 scores for each of the 6 domains are summarised in Supplementary Table 4 and the breakdown of individual domain scores for round 3 are presented in Table 1. The overall consensus was achieved with the mean scores ≥5.25 out of 7 (≥75%) for all items in the 6 domains, three Delphi rounds were sufficient to reach this consensus.

### Guideline Appraisal

The countries from which the guidelines were selected for appraisal based on the GNI per capita, with their corresponding confirmed cases and total deaths and transmission classification based on WHO situation report[1] are summarised in Supplementary Table 5. The comparison of all guidelines, except WHO is summarized in Table 1. There was a significant difference in scores for Domain 4 (p=0.047). The Kruskal Wallis H test p-values for the five reviewers grading G24 (WHO) were 0.51, 0.25, 0.63 and 0.88 respectively, while domains 5 and 6 were significantly different (p=0.009 and p=0.002 respectively). There was good general correlation across all Domains. Monotonicity analysis also revealed scores for the assessor pairs either rising or falling in unison. Finally, inter-rater agreement was excellent using Cronbach’s alpha (table 2).

Likert scores by domain for the national guidelines (G1 to G23) stratified based on the GNI are illustrated in Figure 2. The median (IQR) scores for the national guidelines, for each domain were as follows: Domain-1 6.0 (5.0, 7.0), Domain-2 5.5 (3.5, 6.5), Domain-3 5.3 (2.5, 6.5), Domain-4 3.3 (1.3, 5.5), Domain-5 2.5 (1.0, 5.5) and Domain-6 1.0 (1.0, 2.5). Domains 1, 2 and 3 scored significantly higher than Domains 4, 5 and 6 (p<0.001) with Domain 6 scoring significantly lower than Domains 1 to 5 (p<0.001). The mean (SD) scores by GNI per capita were as follows: high-income 4.10 (2.48); upper-middle income 3.91 (2.14); lower-middle income 2.85 (2.08) and low-income 4.07 (2.21). The overall mean Likert scores by GNI per capita for G1-23, was 3.80. The Likert scores for the high-, upper-middle- and low-income groups were not significantly different whilst the scores for the lower middle-income group were significantly lower than the other three groups (p<0.001, Figure 3).

**Figure 2.**
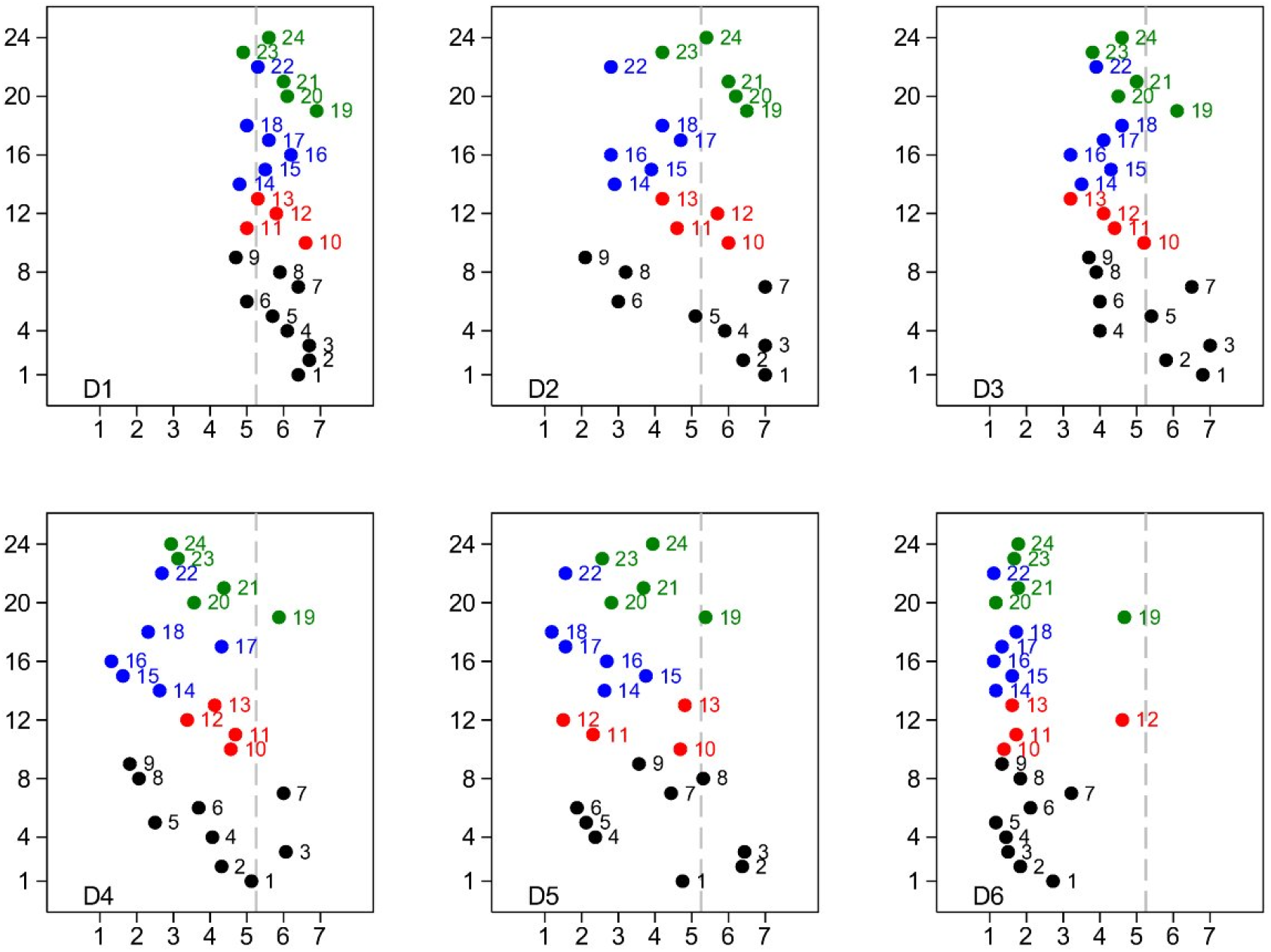
Median Likert scores by Domain (National Guidelines 1 to 24 inclusive) Black - High income; Red - High middle income; Blue - Low middle income; Green - Low income. Domains D1 to D6 are identified on each panel. The vertical line at 5.25 indicates the median score for all Guidelines.

**Figure 3:**
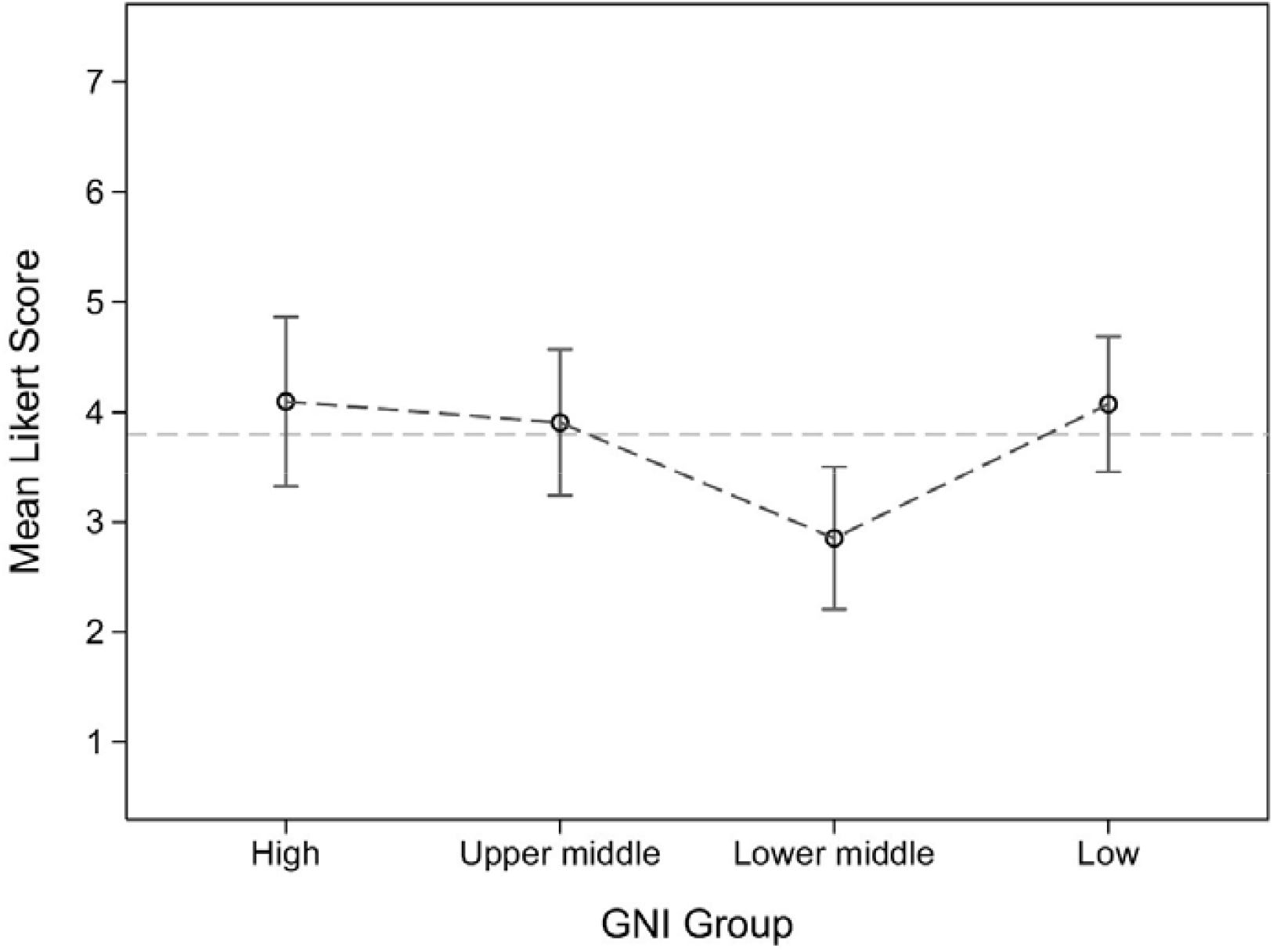
Likert scores by GNI (National Guidelines G1 to G23 inclusive).

Likert scores by domain for the specialty-specific and procedure-specific guidelines (25 to 40) are illustrated in Figure 4. The median scores (IQR) for the professional society and procedural guidelines, for each Domain were as follows: Domain-1 6.0 (5.0, 6.5), Domain-2 5.0 (2.5, 6.5), Domain-3 3.5 (1.0, 6.0), Domain-4 2.5 (1.0, 5.5), Domain-5 1.0 (1.0, 4.0) and Domain-6 1.0 (1.0, 2.0). Domains 1 and 2 scored significantly higher Domains 3 to 6 (p<0.001) whilst Domain 6 was scoring significantly lower than Domains 1 to 5 (p<0.001).

**Figure 4:**
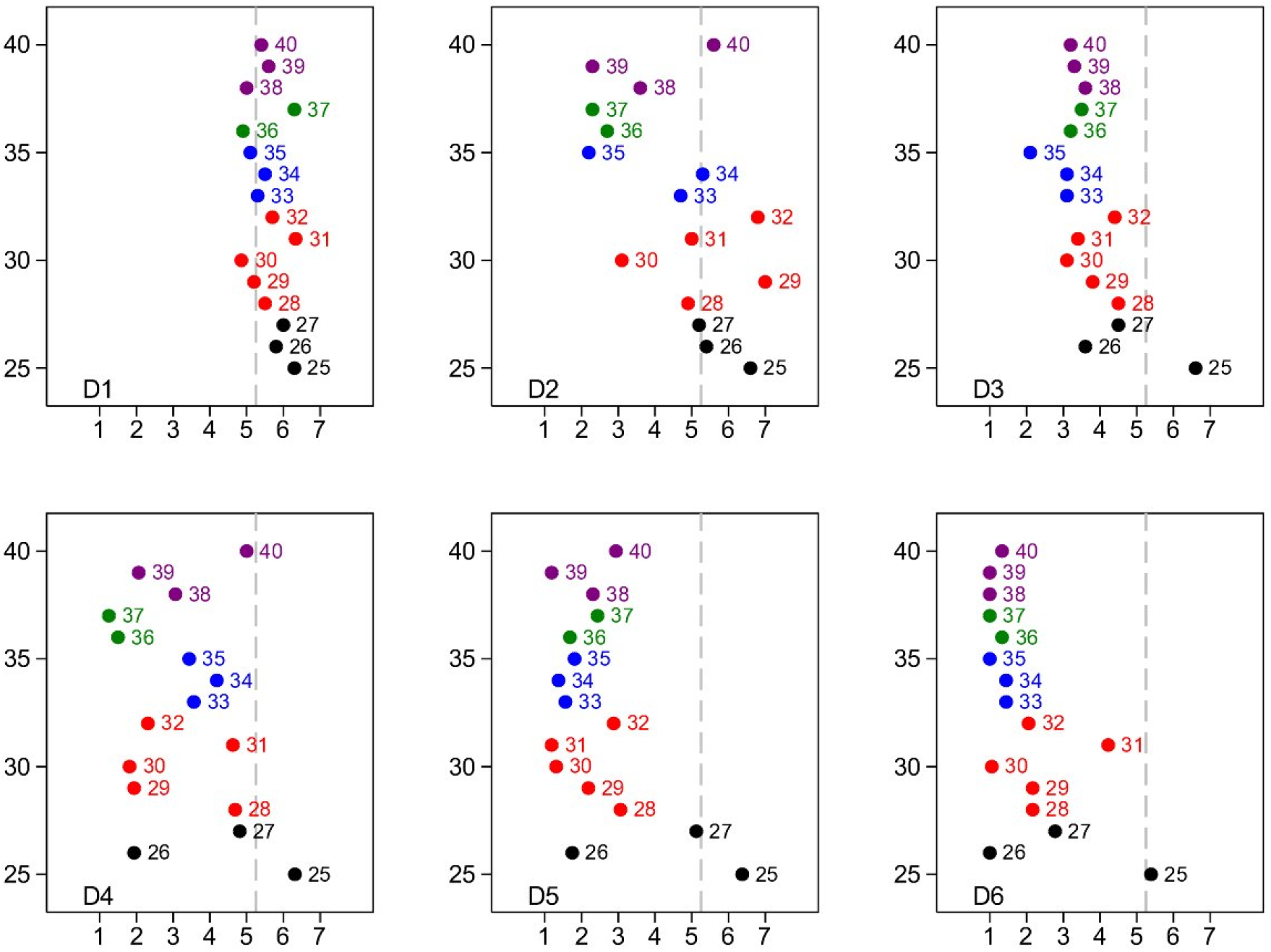
Median Likert scores by Domain (Societal Guidelines 25 to 40 inclusive). Black -Intensive Care Unit society guidelines; Red - Anaesthesia society guidelines; Blue - Intubation procedural guidelines; Green - CPR procedural guidelines; Purple - Handling of deceased guidelines. Domains D1 to D6 are identified on each panel. The vertical line at 5.25 indicates the median score for all Guidelines.

## DISCUSSION

We developed and validated a PPE guideline appraisal tool utilising a validated Delphi consensus building process [19]. The diverse multidisciplinary Delphi panel allowed for the development of a robust generalisable tool. By providing the numerical Likert scale and a high score consensus for the questions to be retained [20, 21], we developed a tool that was objective. Consensus was achieved across all domains despite the diversity of the panel making both the process and tool robust. Discussion and exploration of differences were an important part of this process and this was served well by the iterative methodology inherent in the Delphi process [16]. The final 6 domains created within the tool through the Delphi process consisted of both general and specific recommendations pertinent to the use of strategies to minimise the risk of HCW infections. The tool was then tested for its reliability and validity by appraising 40 varying international/national, specialty-specific and procedure-specific guidelines. Being the first of its kind, there were no pe-existing tools for comparison and to assess for the general validity (construct) of this measurement tool [22]. Involving a diverse expert panel, with a wide range of expertise enhanced its content validity [22]. A Cronbach’s α score of >0.70 demonstrated very good internal construct validity for using this appraisal tool. Sufficient details, presented as supplementary material, can be used to replicate this tool and be evaluated independently.

A range of published international and national infection control guidelines were utilised for validation of the tool. The WHO guidelines have been widely adopted by many low- and middle-income countries that have not published dedicated national guidelines and were an obvious inclusion. Similarly, including published guidelines from countries with different GNI strata added to the strength of the study. Infection control practices and hence guidelines may significantly vary between countries based on socioeconomic status and available resources. In addition, including appraising guidelines from specialty-specific and procedure-specific guidelines was relevant to ensure these guidelines provided engineering, administrative and PPE specific recommendations to HCWs at risk.

The appraisal tool performed well across all domains. All included guidelines scored relatively better in domains 1-3 compared with domains 4-6. Domain-1 of the appraisal tool focused on the ‘demographic’ of any guideline (i.e., whether it was specific to COVID-19, whether it targeted relevant HCWs and if it was based on robust evidence or expert consensus, length and layout and ease of reading/interpreting the guideline). This is important because, there is certainly an information overload and oversupply of guidelines. Domain-2 focused on the recommendations on engineering solutions that are fundamental to infection control practices. This is especially important given that aerosol and fomite transmission of SARS-CoV-2 is plausible, since the virus can remain viable and infectious in aerosols for hours and on surfaces up to days [23]. Domain-3 was PPE-specific that were required for appropriate HCWs protection in different clinical situations. Domains 4-6 were based on recommendations pertaining to administrative solutions for infection control that included PPE training, fit testing of masks, efficient PPE use, PPE reuse, PPE disposal, etc. In particular, domain-6 included recommendations that promote a culture of staff safety, risk and adverse event reporting and staff support. Surprisingly, this domain was the most neglected in most guidelines. We postulate that it could probably due to lack of the resources, incentives and facilities that can be provided to HCW during a surge. It may also be the lack of advocacy on behalf of HCWs. Equally, culture of safety is built over time and ideally should be embedded in clinical practice even outside a pandemic and may not be implemented de novo at the height of the pandemic. Infection prevention goes beyond use of PPE. While PPE provides immediate physical and psychological safety to the HCWs, engineering and administrative solutions help build enduring culture of safety in health systems. Therefore, improvements in these domains is critical to the well-being of the HCWs. None of the national guidelines had a score >5.25 in any of the studied domains. This finding was no different in specialty-specific and procedure-specific guidelines, with only one guideline discussing this in adequate detail. Therefore, we identify domains 4-6 as areas where significant gains can be made across all guidelines assessed.

Our primary intention was not to compare the guidelines, but to validate the guideline appraisal tool. In addition to its utility as a tool for PPE guideline assessment, we believe, this tool can provide a reliable framework while writing new guidelines. Although, guidelines evolve over time as new evidence becomes available, the domains listed in this tool that are based on key principle of infection control are likely to stay relevant. More research is needed to explore the relationship between quality of infection control guidelines and HCW infection risks. A well written guideline that performs well applying our appraisal tool can still be poorly implemented on the ground. Thus, quality of guideline may in itself not guarantee safety and this calls for development of quality metrics that helps track gaps in implementation of guidelines, quality of infection control practices including PPE and HCW infection rates over time.

It should be noted that the two assessors who appraised each guideline were not blinded. This was not necessary as the primary objective was validation of the tool rather than ranking of published guidelines. With excellent inter-rater agreement, there was no need for more assessors. Some guidelines were not in English and Google translator was used. However, Google translator uses a robust and efficient technology with scientific validity [24, 25].

## Conclusion

We developed and validated guideline appraisal tool using a rigorous process. The tool performed well when applied to across several international, national, specialty-specific and procedure-specific guidelines and helps identify the strengths and weaknesses of each guideline. This tool may enable health systems to select and adopt high quality guidelines while optimizing infection control practices and PPE use. A guideline that performs well, when appraised by our tool will not only provide meaningful recommendations for infection prevention in HCWs, but also will help build an enduring culture of staff safety in health systems. More research is needed to explore the gaps in guideline implementation and to explore the relationship between quality of infection control guidelines and HCW infection risks during a pandemic.

## Data Availability

This study does not contain patient information. This study is about developing and validating a guideline appraisal tool for infection prevention in COVID-19

## Acknowledgements

The investigators would like to acknowledge the time and effort volunteered by the following Delphi panel members - Ms Karen Wheeler, Ms Cheryl Baker, Dr Peter Brown, Dr Abdul Samad Ansari, Dr Maximilian Moser, Mr Brendon Gardner, Dr Richard la Nauze, Ms Emma Ridley, Dr Kishan Ajjampur, Ms Rachel Longhurst, Dr Bhagya Ratna Tekula, Ms Deborah Sharp, Mr Reuben Weare, Prof John Botha, Ms Toni Moylan, Mr Jino Jacob, A/Prof Vikas Wadhwa, Mr Gareth James, Dr Kate Lim, Ms Bec Gray, Ms Leah Adams, Ms Simone Redpath, Ms Wendy Brooks, Dr Robert Dawson, Ms. Kellie Bamberry, Ms Laurel Walker, Ms Joanne Stewart, Dr Markus Renner, Ms Amy Brown, Dr Yogesh Apte, Dr Rafatullah Parkar, Mr Long (Matt) Huynh, Dr Gary Braun, Ms Joanne Molloy, Ms Navya Gullapalli, Ms Elena Allen, Dr Prakash Nayagam, Prof Ravindranath Tiruvoipati, Dr Sandeep Kumar Mondal, Ms Shae Whyte-Clarkson, Dr Amit Das, Dr Sourav Dhara, Dr Rana Ray, A/Prof Andrew Davies, Ms Marina Lander, Dr Oles Yehorov, Ms Maria Camona, Dr Elisabeth Nye, Dr Charles Aitken, Dr Ian Young, and Dr Wai Tat Wong. Dr Kiran Shekar acknowledges research support from the Metro North Hospital and Health service.

## Contributors

KS and AS conceived the project idea. The Delphi steering committee comprised of AS, MR, UK, AZ, ZL and KS. The literature review was conducted by AS, MR, AZ, ZL, SM, UK and KS to create the domains and items for Delphi round 1. AS, MS, AZ, MR, UK, SK and ES distributed the Delphi round 2 and 3 surveys and discussed results to reduce and refine the final tool. The 40 guidelines were randomly allocated to the reviewers by AZ. These guidelines were reviewed by AS, AZ, AR, JH, JL, MR, SM, UK, and ZL. The results were collated by AZ and AS. CA conducted the statistical analysis. AS and KS analysed the data and wrote the initial drafts of the manuscript with inputs from MR, AZ and UK. AS, AZ, CA and SM created the tables and figures. AS and KS finalized the manuscript. All authors critically reviewed the manuscript and approved the final version prior to submission.

## Data sharing

All data relevant to this study is provided in the article and in the supplementary material.

